# Creating virtual H&E images using samples imaged on a commercial CODEX platform

**DOI:** 10.1101/2021.02.05.21249150

**Authors:** Paul D. Simonson, Xiaobing Ren, Jonathan R. Fromm

## Abstract

Multiparametric fluorescence imaging via CODEX allows the simultaneous imaging of many biomarkers in a single tissue section. While the digital fluorescence data thus obtained can provide highly specific characterizations of individual cells and microenvironments, the images obtained are different from those usually interpreted by pathologists (i.e., H&E slides and DAB-stained immunohistochemistry slides). Having the fluorescence data plus co-registered H&E or similar data could facilitate adoption of multiparametric imaging into regular workflows, as well as facilitate the transfer of algorithms and machine learning previous developed around H&E slides. Since commercial CODEX instruments do not produce H&E-like images by themselves, we developed a staining protocol and associated image processing to make “virtual H&E” images that can be incorporated into the CODEX workflow. While there are many ways to achieve virtual H&E images, including use of a fluorescent nuclear stain and tissue autofluorescence to simulate eosin staining, we opted to combine fluorescent nuclear staining (via DAPI) with actual eosin staining. We also output images derived from fluorescent nuclear staining and autofluorescence images for additional evaluation.

## INTRODUCTION

Highly multiparametric fluorescence imaging is gaining increased use in research laboratories, particularly those involved in tumor microenvironments and related immunological research wherein the accurate co-localization of many markers within individual and adjacent cells is important [1-3]. CODEX imaging [3,4] is available as a commercial system from Akoya and is one such multiparametric imaging system. Some academic pathology labs, and in particular, hematopathology divisions, are also taking notice, with the possibility of translation to clinical use becoming more likely as the methods are further developed. Pathologists, however, are trained to interpret histology using primarily hematoxylin and eosin (H&E) stained slides and immunohistochemically stained slides using 3,3⍰-diaminobenzidine (DAB) and hematoxylin counterstain. Given the critical importance of proper interpretation of histology for clinical care, pathologists are understandably hesitant when asked to interpret histology using unfamiliar staining and imaging approaches. As machine learning algorithms for automated image interpretation in pathology have thus far also focused primarily on H&E-stained slides, transfer of some of that learning is desirable and could be facilitated by co-registered H&E staining in addition to the multiparametric biomarker data.

In order to incorporate multiparametric fluorescence imaging in clinical use, it is therefore desirable to, in addition to the fluorescence images, have H&E-stained images to compare with fluorescence images [5]. As our work involves a commercial CODEX instrument, we had hoped to perform regular H&E staining on the same tissue used for fluorescence imaging. In the CODEX system, tissue is attached to a glass coverslip rather than a microscope slide. The coverslip is sandwiched between two gaskets (or between a gasket and microscope stage adapter in the updated version) and serves as the bottom of a well, through which fluids are passed during the cyclical staining and imaging process that makes highly multiparametric imaging possible. A coverslip is required since the tissue is imaged on an inverted microscope, and microscope slides would be too thick for the optical objectives used in the usual system. In our experiments, half or more of the coverslips cracked in the process of removing them from the gaskets. Other investigators have reported staining and imaging the coverslips with eosin at the end of the regular CODEX imaging process while the coverslips are still in place (i.e., still between the gaskets).[3] However, their report lacked further details regarding their approach.

We therefore developed our own protocol for creating images using CODEX coverslips that closely resemble traditional H&E-stained tissue sections (“virtual H&E” images, see for examples [5-11]). Similar to another report [3], we leave the coverslip in place on the instrument and use the fluid well created by the coverslip, gaskets, and coverslip holder to stain with eosin. Since eosin is fluorescent and compatible with our Cy3 filter set [12], it is simple to image within the existing system. Unfortunately, hematoxylin does not have similar properties. However, since hematoxylin is primarily used to stain the nuclei, we felt that substituting 4⍰,6-diamidino-2-phenylindole (DAPI) staining for hematoxylin would be sufficient for our purposes (and compatible with our existing DAPI filter set). We then apply relatively straightforward mathematical transformations, similar to those reported elsewhere [5,8,13], to the fluorescence images of the eosin and DAPI staining to create our virtual H&E images.

While 20x magnification is generally sufficient for identifying markers expressed by cells and is the default used by most CODEX users, we found the virtual H&E images somewhat wanting in detail at 20x (Keyence BZ-X800 microscope with Nikon 20x PlanApo lambda NA 0.75 objective), particularly with respect to chromatin patterns for interpretation of hematologic cells. We therefore used a 40x objective (Nikon 40x PlanApo lambda NA 0.95) for virtual H&E imaging.

## METHODS

### Institutional oversight

All methods were carried out in accordance with relevant guidelines, regulations, and approval by the institutional review board of the University of Washington (IRB #43066).

### Eosin and DAPI staining and imaging protocol

An 8.2% eosin Y working solution was prepared using 390 ml 95% ethanol, 50 ml 1% eosin Y, 5 ml 1% phloxine, 2 ml glacial acetic acid, 300 ml of which was then mixed with 100 ml water and 2 ml glacial acetic acid to arrive at the final eosin Y working solution. A 50% solution of ethanol diluted in purified water was also prepared prior to staining. After completion of a normal CODEX imaging run and with the coverslip still in place on the microscope stage, the solution within the coverslip well was removed and replaced 3 times with 700 µl of 50% ethanol in water, being careful to pipet gently so as to not cause tissue to separate from the glass coverslip. This was then replaced with 500 µl of 50% ethanol solution with 0.25 µl of DAPI solution (Akoya Nuclear Reagent; Akoya cat. no. 7000003) and 0.5 µl of eosin working solution added. After incubating for 5 minutes, the solution within the well was then replaced three times with 500 µl of 50% ethanol solution. The well solution was then replaced by CODEX buffer (Akoya cat no. 7000001, diluted 10x in water) and imaged soon thereafter on a Keyence BZ- X800 microscope using DAPI and Cy3 filter sets (Chroma cat. no. 49000 and Chroma cat. no. 49004, respectively) and 0.75 NA Nikon 20x and 0.95 NA Nikon 40x PlanApo lambda objectives.

We found that the staining intensity of eosin was somewhat variable from day to day. If the eosin staining was found to be too weak, the staining process was repeated but with a doubled volume of eosin working solution. The staining intensity was deemed too weak if there was obviously decreased intensity in a tissue scanning pattern prior to imaging the eosin (attributed to photobleaching of autofluorescent molecules, while the eosin signal is expected to be stronger than the autofluorescence for sufficiently stained tissue, resulting in elimination of a photobleaching pattern). If the eosin staining was found to be too strong, the tissue was cleared using the CODEX staining platform, and the staining was repeated with a lower concentration of working eosin solution.

### Conversion of raw fluorescence images to virtual H&E images

Fluorescence images demonstrate signal intensity that is generally linear with respect to the number of fluorescent molecules present, while normal bright field imaging of true H&E-stained slides is essentially a measurement of light absorbance by the dye molecules and follows a logarithmic relationship with respect to concentration of dye molecules [13]. We therefore modeled the conversion of fluorescence images to absorbance images using the following equations

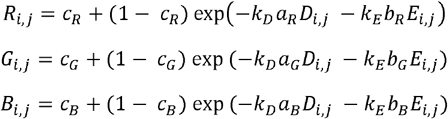

where R, G, and B correspond to the red, green, and blue channels of the resulting virtual H&E image; i and j are pixel indices; **c** values correspond to a postulated expected minimum amount of light (out of focus light, etc.) that is transmitted for a maximally stained specimen for the given channels; **a** and **b** vectors correspond to the expected decrease in bright field light intensity for increasing brightness seen by fluorescence microscopy for DAPI and eosin staining, respectively; *k*_*D*_ and *k*_*E*_ are scalars that are added for convenience for adjusting impact on image output based on DAPI and eosin input images, respectively, and to avoid modifying component values of **a** and **b**; D is the DAPI monochromatic fluorescence image; and E is the monochromatic eosin fluorescence image. Constants were adjusted until there was satisfactory correspondence with corresponding H&E images. The resulting RGB channels form a final virtual H&E image. By omitting the eosin image (E), a virtual hematoxylin-only stained image is seen, similar to what is used for conventional immunohistochemistry. Software for the conversion was developed in python using a Jupyter notebook and using a minimal set of modules that included numpy, pandas, tifffile, and matplotlib.

In addition to conversion of images obtained via eosin and DAPI combination staining, image conversion was also performed on DAPI and autofluorescence images. This was performed for comparison with the DAPI/eosin staining images. The DAPI/autofluorescence images were collected using an Atto 550 filter set (Cy3 filter set) on the first cycle of the regular CODEX imaging (when autofluorescence is expected to be highest) with equivalent mathematical transformations, albeit with different chosen constants *k*_*D*_ and *k*_*E*_.

## RESULTS

Virtual H&E images were considered qualitatively good surrogates for regular H&E-stained images, as determined by two board-certified pathologists (PDS and JRF), very similar to that expected by regular H&E staining. Example images can be seen in Figs. 1-3. The protocol was time sensitive, with gradual loss of eosin staining over time as it dissociated from the tissue specimens and increased generalized background fluorescence from the imaging buffer. Hence, imaging was performed directly after completion of the staining protocol. After imaging with eosin and DAPI, we experimented with tissue clearing by the CODEX instrument, followed by re-imaging. Re-imaging demonstrated that virtually all of the eosin was removed by the CODEX tissue clearing cycle.

**Figure 1.**
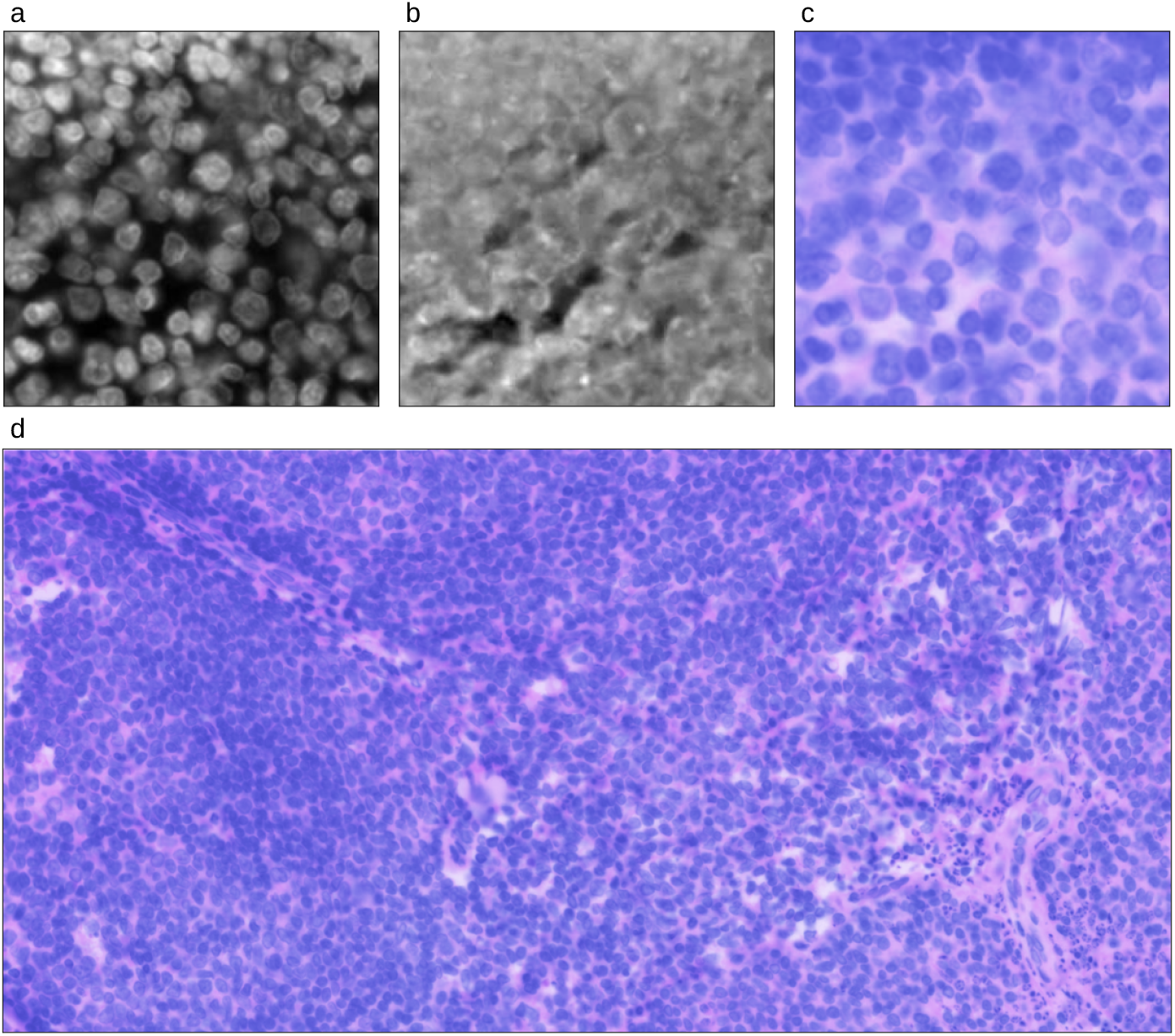
Virtual H&E imaging of a section of tonsil imaged using 40x magnification. (a) DAPI fluorescence image. (b) Eosin fluorescence image. (c, d) Virtual H&E bright field image.

**Figure 2.**
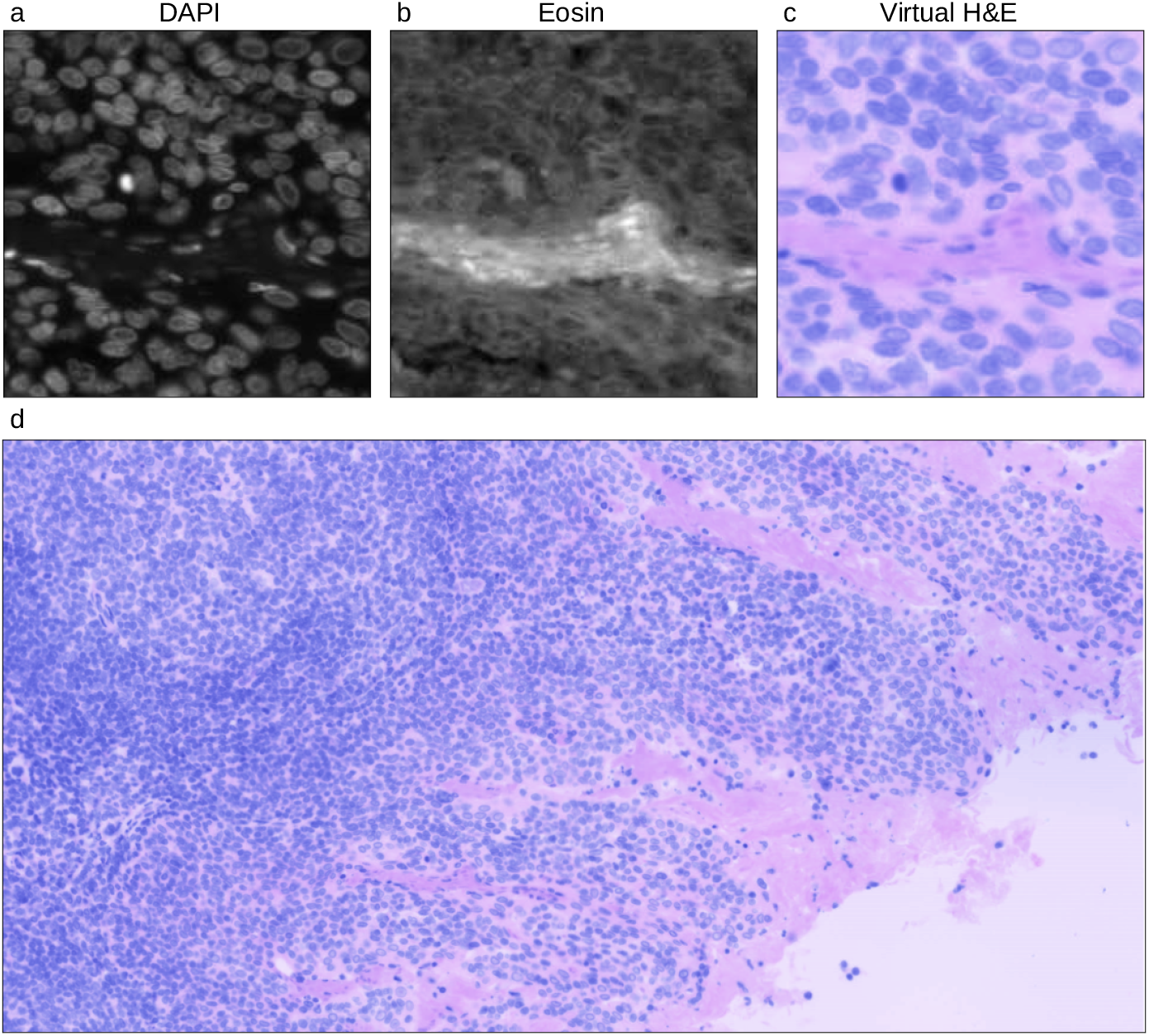
Second example virtual H&E imaging of a tonsil, 40x magnification. (a) DAPI fluorescence image. (b) Eosin fluorescence image. (c, d) Virtual H&E bright field image.

**Figure 3.**
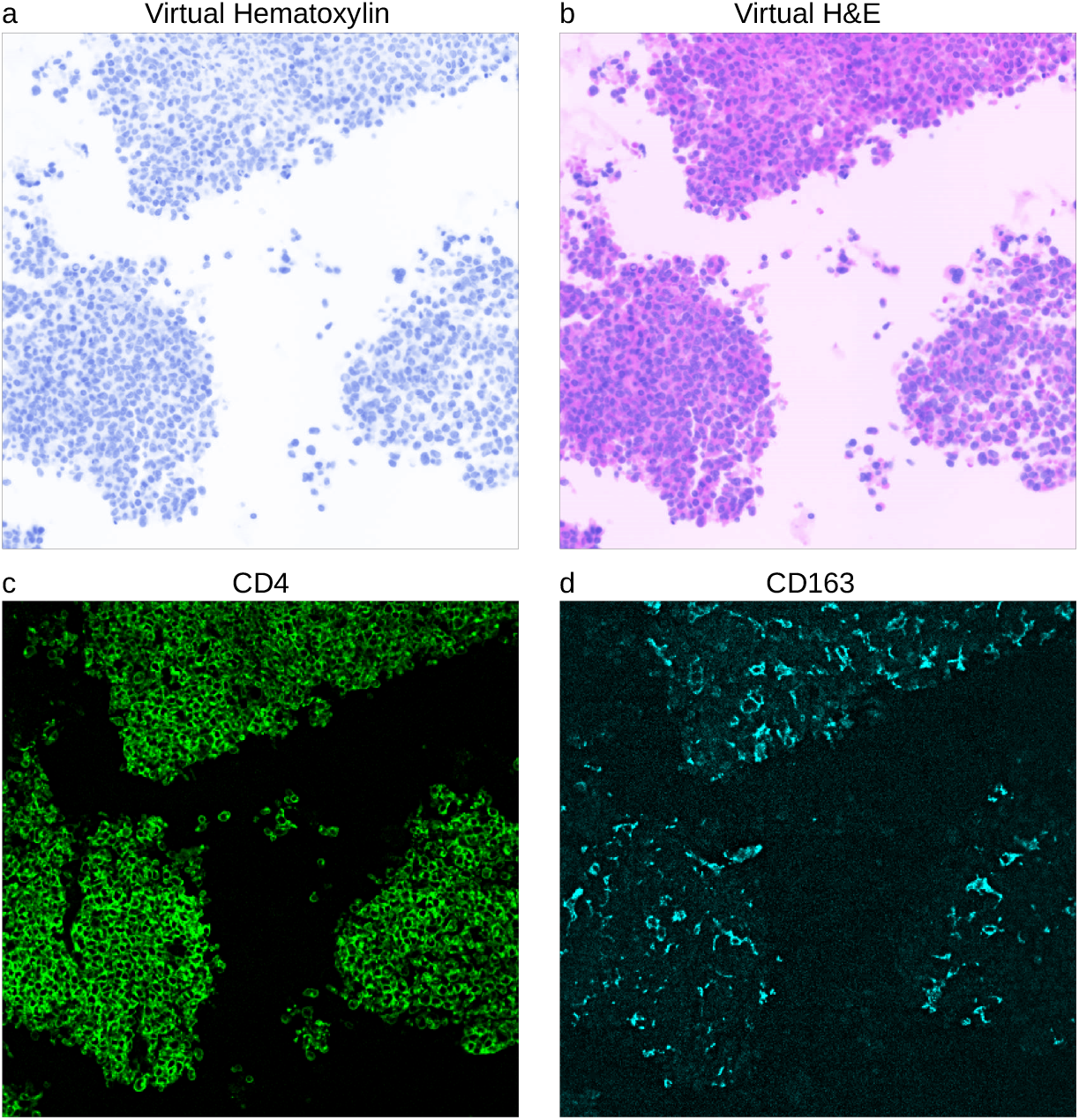
Virtual H&E imaging of a particle preparation of acute monocytic leukemia aspirate. (a) Virtual hematoxylin image (achieved simply by setting *k*_*2*_ = 0), 40x magnification. (b) Virtual H&E image, 40x magnification. (c, d) CD4 and CD163 CODEX fluorescence images, respectively, obtained at 20x magnification.

Virtual H&E images were also created using images collected during the course of normal CODEX imaging, with the DAPI and autofluorescence (captured through an Atto550 filter set) images from the first cycle of imaging used to create the virtual H&E images. While the DAPI images were qualitatively very similar to those captured in our post imaging DAPI-eosin staining protocol, the autofluorescence images showed significant raster scanning photobleaching artifacts that were evident in the final images, more than that seen for eosin staining. See example in Fig. 4.

**Figure 4.**
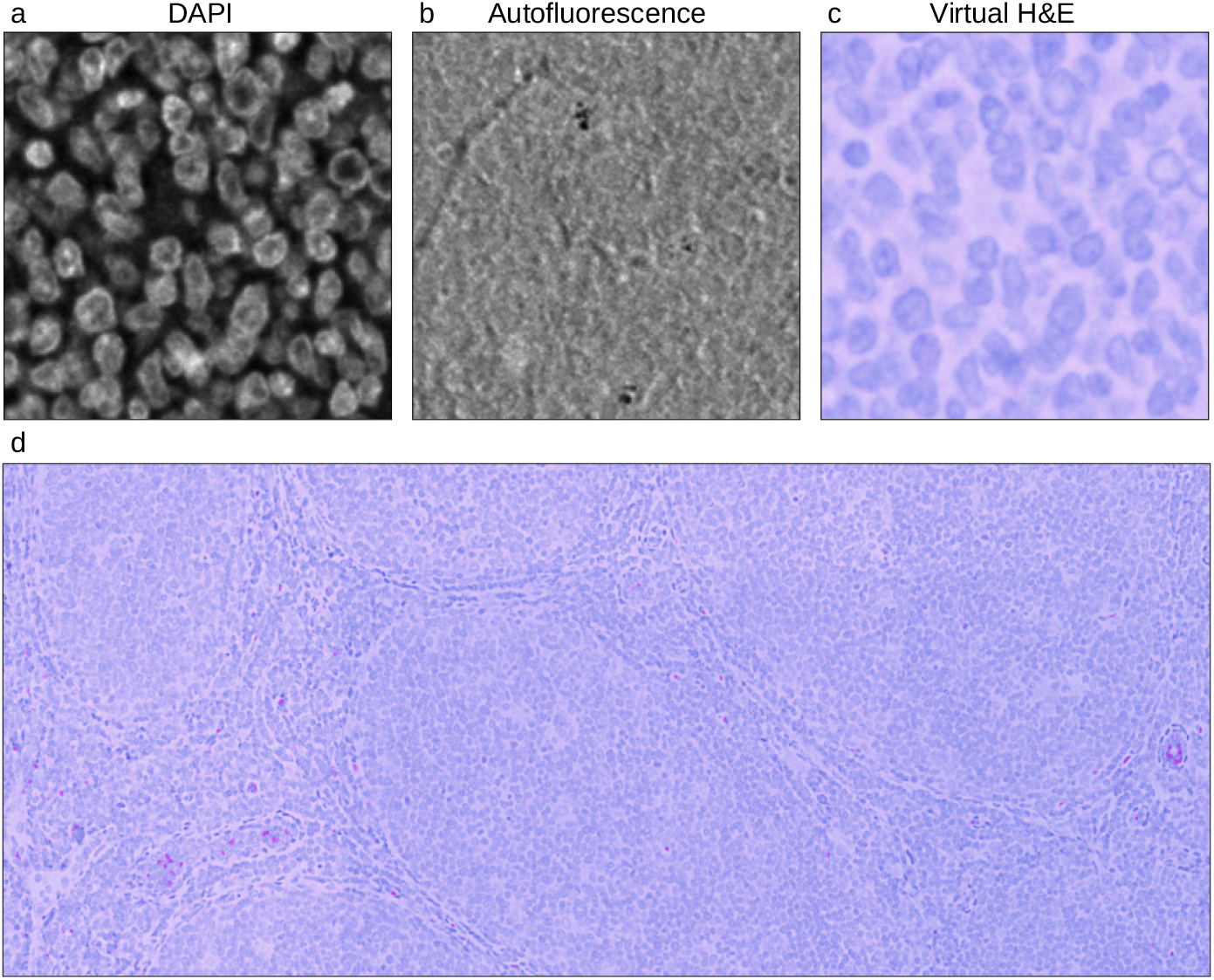
Virtual H&E images of a tonsil created using autofluorescence image instead of eosin fluorescence image, 20x magnification. (a-c) DAPI fluorescence, eosin fluorescence, and virtual H&E bright field images, respectively. (d) Photobleaching of the autofluorescence signal contributes to artifacts due to raster scanning often being present in final H&E images.

## DISCUSSION

As bright field H&E staining is predicted to remain the gold standard by which pathologists are trained to interpret histology images, presenting imaging data acquired via other methods in a virtual H&E representation is desirable[5-10,13]. Here we have presented our method for use with a commercial CODEX imaging system[3,4] that allows us to use the same tissue section to create a virtual H&E image for direct comparison with the fluorescence imaging of many tissue markers. A similar process could be (and is often) followed for other new imaging systems when the samples cannot be (or cannot conveniently be) stained using H&E and bright field imaging.

There are multiple methods that could be applied to create virtual H&E images from tissue imaged on a CODEX system. In addition to the method we have presented, another option includes using images of the tissue autofluorescence (collected, for example, using a GFP or Atto 550 filter set) for virtual eosin staining and DAPI fluorescence for virtual hematoxylin staining [5]. This approach eliminates manual staining at the risk of a mismatch between the autofluorescence signal and what would be the true eosin staining pattern. Despite this risk, for many applications the difference will be small and not affect tissue interpretation. Nevertheless, we determined that actual eosin staining would be preferable, which prompted the development of a staining procedure and associated software. When both imaging approaches were directly compared, we found the eosin-stained imaging to be of higher quality, primarily due to the strong effects of photobleaching seen in the autofluorescence images, which were less pronounced in the eosin images (which is to be expected given the generally short photobleaching lifetime for most autofluorescence).

While imaging at 20x allows for easy co-registration of virtual H&E images and fluorescence staining of biomarkers, co-registering 20x images with ones collected at 40x is less convenient. Therefore, users may consider capturing fluorescence images for conversion to virtual H&E images using both 20x and 40x magnifications. Down-sampling of 40x images with alignment based on the raw DAPI fluorescence images is also a viable option when deemed necessary for co-registering with fluorescence data of other markers collected at 20x magnification.

Given that the transformation to virtual H&E images involves applying exponential-decay transforms, some image detail can actually appear to be lost after transformation, which was particularly noticed in comparing transformed and non-transformed images of cell nuclei. By making the option available to switch between exponential-transformed and linear-transformation images, it is likely that pathologists will quickly adapt to and possibly even prefer having the non-transformed DAPI images, despite being less similar to true H&E images.

An important limitation of this approach is the lack of visualization or altered visualization of pigments present in the specimen since these will be displayed as some mix of hematoxylin and eosin coloring. Examples include hemoglobin (appearing as magenta in color rather than red) and melanin. For evaluation of pigments intrinsic to the sample, additional tissue sections with traditional H&E staining will likely be required. Also, as eosin might have different fluorescence properties in different molecular environments (e.g., shifted emission spectrum when bound to protein, self-quenching effects for high molecule densities, etc.), we also recognize that, even with using the same molecule, the correlation between bright field and fluorescence imaging may be more complex than our simple transformation allows, and more sophisticated transformations (e.g., non-linear or deep learning GAN models) might ultimately yield better virtual H&E images. Finally, the images produced via fluorescence are a combination of eosin fluorescence and autofluorescence, so care must be taken to ensure sufficiently strong eosin staining is present to ensure the autofluorescence contribution is negligible.

In summary, we have presented a straightforward staining protocol and analysis algorithm that should be of interest to investigators who are using CODEX systems and wish to create virtual H&E images of the same tissue section used for imaging biomarkers. The images will be helpful for trained pathologists who are asked to interpret the images, whether for research questions or eventual clinical use.

## Data Availability

Relevant python code and example data files will be made available at github.com/SimonsonLab/VirtualHE.

## SUPPLEMENTARY MATERIAL

Updated source code and associated example data are available for download upon acceptance of this manuscript for publication.

## References

1. de Vries NL, Mahfouz A, Koning F, et al. Unraveling the Complexity of the Cancer Microenvironment With Multidimensional Genomic and Cytometric Technologies. Front Oncol 2020; 10: 1254.

2. Hartmann FJ, Bendall SC. Immune monitoring using mass cytometry and related high-dimensional imaging approaches. Nat Rev Rheumatol 2020; 16: 87–99.

3. Schurch CM, Bhate SS, Barlow GL, et al. Coordinated Cellular Neighborhoods Orchestrate Antitumoral Immunity at the Colorectal Cancer Invasive Front. Cell 2020; 183: 838.

4. Goltsev Y, Samusik N, Kennedy-Darling J, et al. Deep Profiling of Mouse Splenic Architecture with CODEX Multiplexed Imaging. Cell 2018; 174: 968–981 e915.

5. Lahiani A, Klaiman E, Grimm O. Enabling Histopathological Annotations on Immunofluorescent Images through Virtualization of Hematoxylin and Eosin. J Pathol Inform 2018; 9: 1.

6. Elfer KN, Sholl AB, Wang M, et al. DRAQ5 and Eosin (‘D&E’) as an Analog to Hematoxylin and Eosin for Rapid Fluorescence Histology of Fresh Tissues. PLoS One 2016; 11: e0165530.

7. Li D, Hui H, Zhang Y, et al. Deep Learning for Virtual Histological Staining of Bright-Field Microscopic Images of Unlabeled Carotid Artery Tissue. Mol Imaging Biol 2020; 22: 1301–1309.

8. Cahill LC, Giacomelli MG, Yoshitake T, et al. Rapid virtual hematoxylin and eosin histology of breast tissue specimens using a compact fluorescence nonlinear microscope. Lab Invest 2018; 98: 150–160.

9. Fereidouni F, Harmany ZT, Tian M, et al. Microscopy with ultraviolet surface excitation for rapid slide-free histology. Nat Biomed Eng 2017; 1: 957–966.

10. Reder NP, Glaser AK, McCarty EF, et al. Open-Top Light-Sheet Microscopy Image Atlas of Prostate Core Needle Biopsies. Arch Pathol Lab Med 2019; 143: 1069–1075.

11. Glaser AK, Reder NP, Chen Y, et al. Light-sheet microscopy for slide-free non-destructive pathology of large clinical specimens. Nat Biomed Eng 2017; 1.

12. Ali H, Ali S, Mazhar M, et al. Eosin fluorescence: A diagnostic tool for quantification of liver injury. Photodiagnosis Photodyn Ther 2017; 19: 37–44.

13. Giacomelli MG, Husvogt L, Vardeh H, et al. Virtual Hematoxylin and Eosin Transillumination Microscopy Using Epi-Fluorescence Imaging. PLoS One 2016; 11: e0159337.

